# Acceptability of AI-applications in routine clinical care for children and adolescents: perspectives of parents and healthcare professionals

**DOI:** 10.64898/2026.09.10.26362703

**Authors:** Feng Lin, Anne C. Bischops, Janna-Lina Kerth, Thomas Meissner, Tommaso Bruni, Simon B. Eickhoff, Bert Heinrichs, Juha M. Lahnakoski, Juergen Dukart

## Abstract

With the rapid advancement of artificial intelligence (AI) technologies, growing interest has emerged in its potential application in pediatric healthcare. Understanding the factors influencing AI acceptability among parents and clinical staff is an important prerequisite for facilitating its integration in clinical routine. To date, these factors have not been systematically assessed across different stakeholder groups. Here we investigated AI acceptability among parents (first cohort n = 198; second cohort: n = 79) and pediatric health care professionals (n = 33) across different disease, diagnosis or treatment scenarios. The effects of demographic variables and contextual predictors, such as data privacy, AI knowledge and perceived disease severity, on willingness to use AI were evaluated. More liberal data privacy was associated with reduced willingness to use AI (*p* < .001). A higher perceived disease severity was linked to higher willingness to use AI (*β = .10, p* = .013) in the second parent group. When AI and clinicians’ recommendations conflicted, parents were more likely to choose AI over clinician’s judgment in treatment compared to diagnosis scenarios. Healthcare professionals showed similar patterns but additionally weighted perceived disease severity when resolving disagreements. These findings highlight the importance of addressing stakeholder concerns regarding disease specific customization, accuracy, data privacy and accountability when introducing AI into pediatric healthcare settings.

**Highlights:**

- Parents are more willing to use AI for conditions they perceive as more severe
- Data sharing beyond institutional level significantly reduces willingness to use AI
- Digital literacy and previous medical knowledge facilitate tolerance for AI errors
- AI applications are preferred in treatment over diagnosis scenarios
- Human judgements are prioritized over AI during inconsistent recommendations

## Background

The integration of artificial intelligence (AI) into clinical care has generated considerable enthusiasm, yet its acceptance among end-users remains uneven and poorly understood (1,2). While some studies report high clinician openness to AI applications in clinical care (3,4), others reveal concerns about accountability, transparency, and bias in AI-assisted medical decisions (5). Among patient populations, acceptance appears context-dependent, varying by disease type, perceived risk, and trust in technology (6,7). This variability suggests that AI adoption in healthcare cannot be understood through universal models of technology acceptance, but rather requires careful attention to the specific clinical contexts, stakeholder roles, and decision making structures in which AI systems are deployed.

Pediatric healthcare presents a particularly complex context for AI implementation. Unlike adult healthcare, its decision making authority is distributed among multiple stakeholders: parents or other guardians who provide consent, clinicians who bear professional responsibility, and in many cases, children themselves (8). This relationship introduces unique tensions around trust, risk tolerance, and the locus of decisional authority (9,10). Furthermore, pediatric diagnoses often involve developmental considerations and lower tolerance for error, potentially amplifying concerns about AI reliability and safety (11). Yet empirical evidence on how parents and pediatric clinicians perceive AI assisted decision making, particularly when their judgments diverge from AI recommendations remains scarce. The question of whether these two understanding stakeholder groups share similar priorities regarding AI accuracy, transparency, and data governance, or whether their acceptance is shaped by different concerns, has received little systematic investigation.

The recent adoption of the EU Artificial Intelligence Act (12) underscores the necessity of user acceptance in medical contexts. By classifying many medical AI systems as high risk and mandating transparency, human oversight, and conformity assessments, the Act implicitly assumes that such measures will enhance trust and adoption. However, whether end users, particularly in shared decision making contexts like pediatric care, weigh transparency against other factors like accuracy, disease severity, or professional judgment, remains an open question.

Despite growing interest in AI acceptance, several knowledge gaps remain. First, most studies focus on either patients or clinicians alone, leaving unclear whether these two groups share similar priorities regarding AI use (9,13). In case of different expectations or concerns about AI from parents and clinicians, implementation efforts that satisfy one group may fail to address the other’s priorities. Second, acceptance is typically measured as a general attitude rather than being tied to specific clinical tasks (e.g., diagnosis vs. treatment recommendation) or contextual variables like disease severity or data sharing arrangements. This obscures whether users’ openness to AI is stable across situations or varies with perceived stakes and systemic factors. Third, the role of individual differences, such as digital literacy, prior AI and medical knowledge, or baseline risk perception, in moderating acceptance has received limited attention, particularly among non-professional stakeholders like parents. Understanding these individual level factors is essential to facilitate informed engagement with AI applications.

To address these gaps, we conducted an online-questionnaire study with parents and pediatric clinicians. We systematically varied clinical application scenarios (diagnosis vs. treatment), AI accuracy, and data-sharing scope. Participants responded to realistic scenarios in which specific AI applications (e.g., AI analysis of medical scans, AI-powered drug delivery, etc., see Table 1) are involved, and answered questions designed to reflect their acceptance to use AI. Participants were presented with varying data privacy levels, acceptable accuracy or error thresholds, and preferences for decision-making when human-AI recommendations conflict. We also assessed individual-level moderators including perceived disease severity, digital usage patterns, prior AI and medical knowledge, and demographic characteristics. By examining acceptance patterns across stakeholder groups and situational contexts, we aim to identify the contextual and individual factors that facilitate or hinder AI application in pediatric care, and to provide empirically grounded insights for the design of AI systems, implementation strategies, and regulatory frameworks in pediatric healthcare settings.

## Methods

### Questionnaire design

In experiment 1,we designed a questionnaire featuring four diseases (pneumonia, developmental delay, asthma, and cancer) and their corresponding AI-application scenarios (e.g., Fig 1B). These diseases were selected based on their prevalence in pediatric populations and their varying clinical severity (see Table 1). The featured AI applications are currently available on the market or in international clinical use (not necessarily in Germany). To ensure accessibility across all education backgrounds, we described core features in simple terms and provided functional examples (see Supplemental materials). Participants answered questions based on the specific background of each disease and its AI application.

In experiment 2, to verify the effect of application scenarios on acceptability found in experiment 1, we expanded the Experiment 1 questionnaire by doubling the number of scenarios (see Table 1) where each disease had AI applications at both the diagnosis and treatment scenarios to exclude potential disease-specific effects. Additionally, we measured further variables for predictive modeling, including previous medical knowledge, perceived disease severity, and previous experience with the mentioned diseases. The detailed study design and variables are visualized in Fig. 1.

**Table 1.**
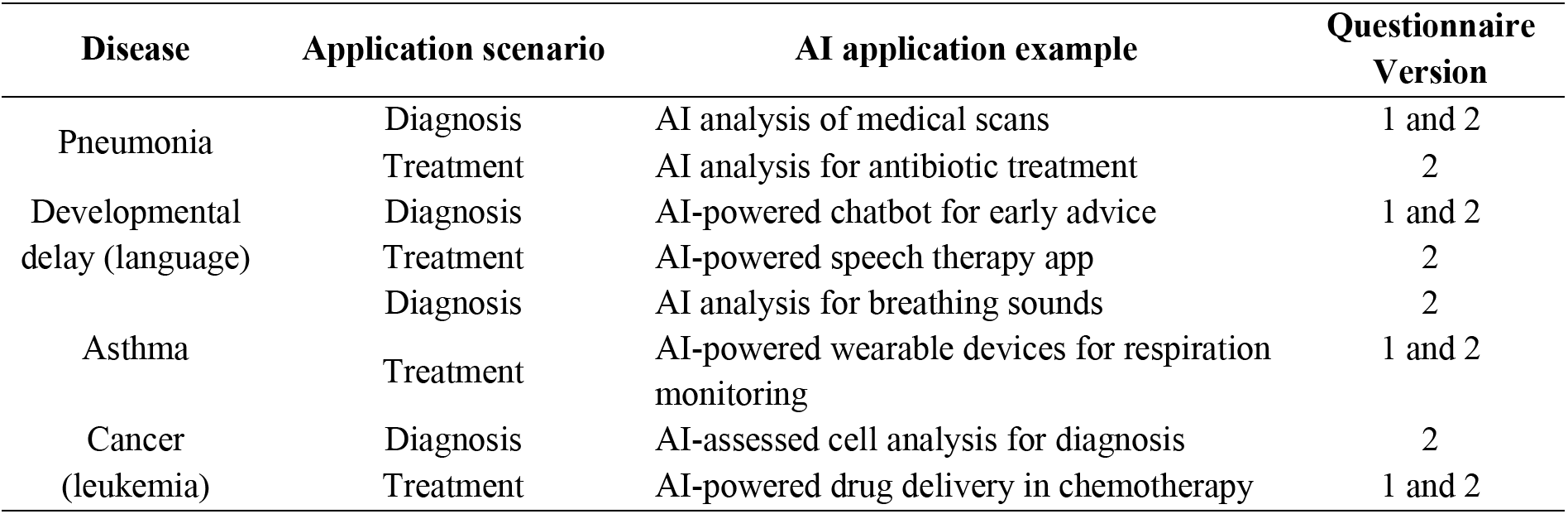
Summary of Diseases Scenarios and Related AI Applications.

**Fig. 1.**
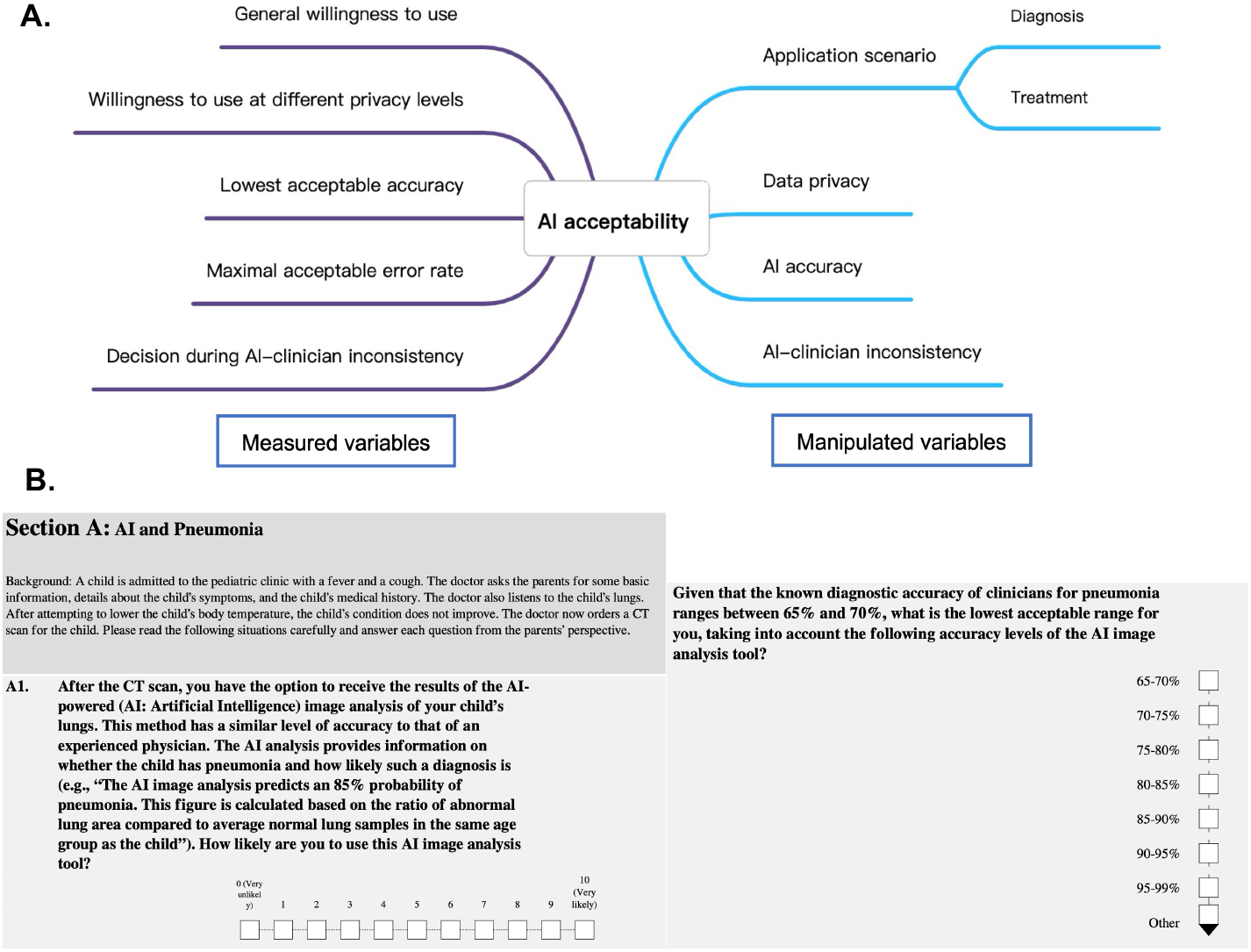
Study design and materials. (**A**) Flowchart of measured and manipulated variables for the questionnaires. (**B**) Example questions for the online questionnaires (full versions of all questionnaires see Supplemental materials).

### Participants and Procedure

Parent participants were recruited anonymously via social media and local advertisements. Eligibility for the parental groups required having at least one child under the age of 18. Clinicians were recruited via social media and professional network advertisements. The eligibility for the clinician group required having experience working with children or adolescents in a medical environment. Participants were asked to complete an online questionnaire-based assessment hosted on LimeSurvey. The study was approved by the ethics committee at the Medical Faculty of Heinrich-Heine-University Düsseldorf (protocol number: #2023-2531), and all participants provided informed consent by selecting a mandatory confirmation box before proceeding. To maintain data integrity, we implemented measures to ensure no participant received the questionnaire link twice.

The final analysis included 198 parents in the first parent group (age: *M* = 37.98, *SD* = 6.69, range 24-69 years, 166 females) after excluding two participants with missing education data, 79 parents in the second parent groups (age: *M* = 38.49, *SD =* 6.48, range 24-54 years, 59 females), and 33 clinicians (age: *M* = 31.94, *SD =* 7.64, range 21-53 years, 28 females). The second parent group was recruited after analysis of the first parent group to replicate and extend the initial findings.

### Data preprocessing

Complete responses were exported from LimeSurvey as separate CSV files for each participant group and encoded individually. Responses to 10-point Likert scale questions were used directly as numeric values without transformation, while accuracy-related responses were calculated based on the difference between the selected range and the provided baseline. For example, if a participant chose a 95–99% accuracy range against an 80% baseline, the answer was converted to a value of 15. Similarly, for maximal error rate questions, the upper limit of the range was recorded as the response. Multiple-choice questions regarding disagreement between AI and clinicians were encoded as categories for the analysis. In the parent groups, these represented trusting and choosing AI, trusting and choosing clinicians, or choosing clinicians or AI when both were deemed possible. These options were adapted for the clinician group to reflect a first-person perspective, where selections represented trusting and choosing their own professional judgment over AI.

### Statistical analysis

All statistical analyses were conducted in R (v4.4.2) (14). Participant age was compared across the three groups using a one-way analysis of variance (ANOVA) after assessing homogeneity of variance. Significant omnibus effects were followed by Tukey-adjusted pairwise comparisons.

To account for repeated measures within subjects, continuous outcome variables were analyzed using linear mixed models (LMMs) via the *lme4* package (15) with random intercepts for participants. Model estimation utilized restricted maximum likelihood (REML) for parameter inference and maximum likelihood (ML) for model comparisons. Significance tests were derived via Satterthwaite’s approximation of denominator degrees of freedom using the *lmerTest* package (16). Across all LMMs, continuous predictors were mean-centered before model fitting to improve interpretability, and the first level of categorical variables served as the reference (e.g., ’female’, ‘diagnosis’, ‘no prior experience’). Estimated marginal means were used for post hoc comparisons of significant interactions. Effect sizes for group comparisons were reported as Hedges’ *g* to reduce small-sample bias. For contrasts derived from linear mixed-effects models, model-adjusted Hedges’ *g* was calculated from the estimated marginal mean difference standardized by the model residual standard deviation. Detailed model specifications, including predictors of interest and covariates for each dependent variable and participant groups are provided in Supplementary Table S1.

For categorical choices in decisions during AI-clinician inconsistency, Bayesian categorical regression models were implemented via the *brms* package (17–19) utilizing Stan (20) with random intercepts per subject. These models were estimated using four Markov chains of 2,000 iterations each (adapt-delta = 0.95), with effects evaluated via 95% credible intervals (CrI). Odds ratios (OR) for numeric predictors (e.g., digital usage score) were calculated by exponentiating the posterior regression coefficients (OR = exp(*β*)), representing the change in the odds associated with a one-unit increase in the predictor. This provided point estimates and corresponding 95% credible intervals were obtained by exponentiating the posterior 95% credible intervals of the regression coefficients.

#### General willingness to use AI

To evaluate general willingness to use AI, separate LMMs were first calculated for each participant group to account for their distinct predictor sets. For the first parent group, fixed effects included AI application scenario (diagnosis vs. treatment) and gender, alongside demographic covariates comprising age, family income level, years of education, maximal child age, digital usage score (i.e., daily count of distinct digital products used), and previous AI knowledge. For the second parent group, the model included all fixed effects and covariates from the first parent group, with the addition of disease experience (yes/no) as a fixed effect, alongside previous medical knowledge and perceived disease severity as covariates. For the clinician group, application scenario and gender remained the primary fixed effects, while covariates included age, working experience (years working with children or adolescents), perceived disease severity, previous AI knowledge, and digital usage score. To assess disease-specific differences in general willingness across participant groups, a LMM model was fitted with disease, group, and their interaction, with application scenario and shared demographic variables (i.e., age, gender, previous AI knowledge and digital usage score) as covariates.

To test the replicability of the initial findings across the parental datasets, an identical LMM utilizing the shared factors (previous AI knowledge, digital usage score, and application scenario) and covariates (age, gender, years of education, income, and maximal child age) between cohorts was fitted to the second parent group. Finally, to test for differences between professional and parental roles, the clinician dataset was merged with the second parent group, and an LMM was conducted utilizing a “group” fixed effect alongside shared predictors (age, gender, previous AI knowledge, digital usage score, application scenario, and perceived disease severity). The interaction between group and digital usage was tested to examine whether the association between digital engagement and willingness to use AI varied between parents and clinicians. This analysis was motivated by evidence that prior technological experience, digital competence, and familiarity with clinical technologies can facilitate healthcare professionals’ acceptance of AI (3,21,22).

#### Lowest acceptable accuracy to use AI

Predictive modeling for accuracy difference (i.e., reported lowest acceptable accuracy − baseline accuracy of the AI applications) followed the exact group-specific structures detailed for general willingness. Within-group analyses involved separate LMMs executed for the first parent group, second parent group, and clinician group using their respective fixed effects and covariates. Cross-sample comparisons were conducted using the same pooling strategy, wherein replicability was assessed by running the baseline shared-predictor LMM on the second parent group, and group differences between parents and clinicians were evaluated by pooling the datasets (Parent Group 2 vs. Clinicians) and including the respective “group” factor as a fixed effect alongside the shared predictors across samples.

#### Maximal acceptable error rate to use AI

Because data collection parameters for the maximal error rate varied by participant group, modeling strategies were adjusted accordingly. For the first parent group, since maximal error rate was only measured during treatment scenarios, the ‘application scenario’ predictor was omitted due to a lack of variable levels. Furthermore, because this metric composed a single observation per subject in this group, a generalized linear model (GLM) with a Gamma distribution and log link was utilized to account for the non-normal distribution of independent observations. Predictors for this GLM included age, gender, years of education, family income level, previous AI knowledge, digital usage score, and maximal child age. In contrast, for the second parent and clinician group where repeated measures were present, standard LMMs were applied using their group-specific fixed effects and covariates. Group comparisons between the second parent group and clinicians, were performed using pooled LMM frameworks restricted to shared predictors and adjusting for ‘group’ fixed effects.

#### Willingness to use AI across privacy levels

To evaluate the impact of data privacy on acceptance, privacy level was added as a critical within-subject fixed effect across all cohorts. For both parent groups and the clinician group, separate LMMs were conducted to predict the dependent variable “willingness to use AI at different privacy levels”. In addition to the standard group-specific main effects and covariates, these models included privacy level as a fixed effect and a primary interaction term between gender and privacy level to test whether the effect of privacy varies by gender. Group comparisons between parents and clinicians were evaluated by fitting the pooled LMM framework (incorporating privacy level and the gender * privacy interaction) alongside the respective ‘group’ fixed effects.

#### Decisions during AI-Clinician inconsistency

To analyze individuals’ decisions when faced with conflicting AI and clinician recommendations, Bayesian categorical regression models were deployed for each participant group. The multi-option response variable used “trust clinicians and choose clinicians” as the reference category. Predictors for these models were identical to those established in the LMMs. Replicability of two parent datasets was tested by running the shared-predictor Bayesian model on the second parent cohort as used in the first parent group. Group comparisons between the second parent group and clinicians were executed by merging the respective data frames and adding a group-level fixed effect to the primary Bayesian *brms* model.

## Results

### Demographic comparison

Age differed significantly across the three participant groups, *F*(2, 307) = 12.57, *p* < .001. Tukey-adjusted post hoc comparisons showed that clinicians were significantly younger than both Parent Group 1 (mean difference = 6.04 years, 95% CI [3.05, 9.03], *p* < .001) and Parent Group 2 (mean difference = 6.55 years, 95% CI [3.26, 9.85], *p* < .001), whereas the two parent groups did not differ significantly in age (mean difference = 0.51 years, 95% CI [−1.60, 2.63], *p* = .835).

### General willingness to use AI

For the first group of parents, the linear mixed-effects model revealed a significant main effect of application scenario (*p* < .001, Hedges’s *g* = 0.67), with higher general willingness to use AI for treatment applications (*M* = 8.04, *SD* = 2.32) than for diagnosis applications (*M* = 6.54, *SD* = 3.00). Males reported a greater overall willingness (*M* = 7.97, *SD* = 2.55) to use AI for their children than females (*M* = 7.15, *SD* = 2.81) (*p* = .046, Hedges’s *g* = 0.32). These effects were not replicated in the second group of parents (see Fig. 2A, B). The willingness scores for diagnosis (*M* = 7.57, *SD* = 2.57) and treatment applications (*M* = 7.88, *SD* = 2.53) were comparable (*p* = .060, Hedges’s *g =* 0.14), and there was no significant effect of gender (*p* = .702, Hedges’s g = 0.10). However, across the full sample, maximal child age was a positive predictor of willingness (*β = .24*, *p* = .019). As shown from the LMM model, perceived disease severity (*β = .10, p* = .013) and maximal child age (*β = .24*, *p* = .024) significantly increased willingness (see Fig. 2C, D). While no significant predictors were identified in the clinician group, a significant group and digital usage interaction was observed (*β = -.26, p* = .043), indicating that the association between digital usage and willingness to use AI was stronger among clinicians than among parents (Fig. 2E). Complete statistical parameters including t-statistics, degree of freedom (*df*), and 95% confidence intervals are detailed in Supplementary Table S2.

A significant disease × group interaction was observed, *F*(6, 1445.36) = 2.34, *p* = .030. Clinicians reported lower willingness than the first parent group for asthma (*p* = .031, Hedges’s *g* = −0.41) and lower willingness than the second parent group for asthma (*p* = .016, Hedges’s *g* = −0.46) and cancer (*p* = .020, Hedges’s *g* = −0.45). No other disease-specific group differences were significant after Holm correction. Detailed means, standard deviations, estimated marginal mean contrasts, *p* values, and model-adjusted Hedges’ *g* values for each disease are reported in Supplementary Table S3.

**Fig. 2.**
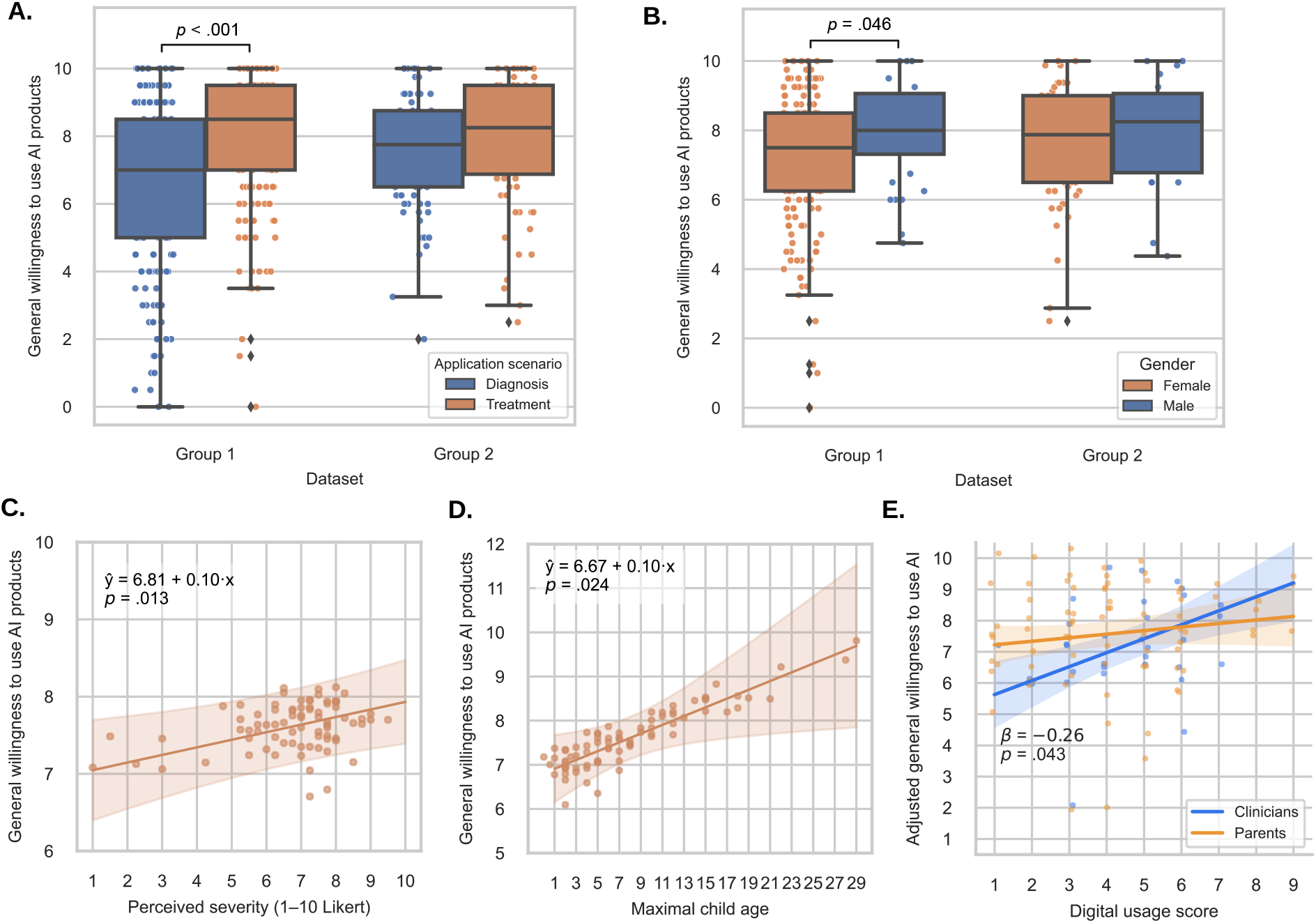
Effects on general willingness to use AI. **(A)** Willingness by application scenario (diagnosis vs. treatment) in the two parent groups. A significant difference was observed only in parent group 1. **(B)** Willingness by gender in the two parent groups. A significant difference was observed only in parent group 1. **(C)** Association between perceived disease severity and willingness to use AI in parent group 2. Points represent participant-level responses adjusted for the remaining covariates in the linear mixed-effects model. The solid line shows the model-predicted relationship, and the shaded area represents the 95% confidence interval (CIs). **(D)** Association between maximal child age and willingness to use AI in parent group 2. Points represent participant-level responses adjusted for the remaining covariates in the linear mixed-effects model. The solid line shows the model-predicted relationship, and the shaded area represents the 95% CIs. **(E)** Interaction between participant group and digital usage on general willingness to use AI. Points represent adjusted participant-level responses. Solid lines show model predictions with 95% confidence intervals.

### Lowest acceptable accuracy to use AI

Perceived disease severity positively predicted the lowest acceptable accuracy across both the second parent group (*β = .24, p* < .001; see Fig. 3B) and the clinician group (*β = .27*, *p* < .001; see Fig. 3C). However, the specific accuracy thresholds varied by application scenario and group. Among the first parent group, participants required a significantly higher accuracy for treatment applications (*M* = 13.13, *SD* = 9.79) than for diagnosis applications (*M* = 9.68, *SD* = 8.64; *p* < .001, Hedges’s *g =* 0.50; Fig. 3A). Conversely, the second parent group showed the opposite pattern, requiring slightly higher accuracy for diagnosis (*M* = 11.60, *SD* = 8.32) than for treatment applications (*M* = 11.04, *SD* = 7.6; *p* = .025, Hedges’s *g =* -0.09).

Finally, when evaluating parent and clinician groups together, the “accuracy gap” (the difference between lowest acceptable accuracy and baseline) was significantly larger among parents (M = 11.41, SD = 8.11) than among clinicians (M = 10.38, SD = 7.24; *p* = .002, Hedges’ *g* = 0.39; Fig. 3D). Complete statistical parameters including t-statistics, degree of freedom (*df*), and 95% confidence intervals are reported in Supplementary Table S4.

**Fig. 3.**
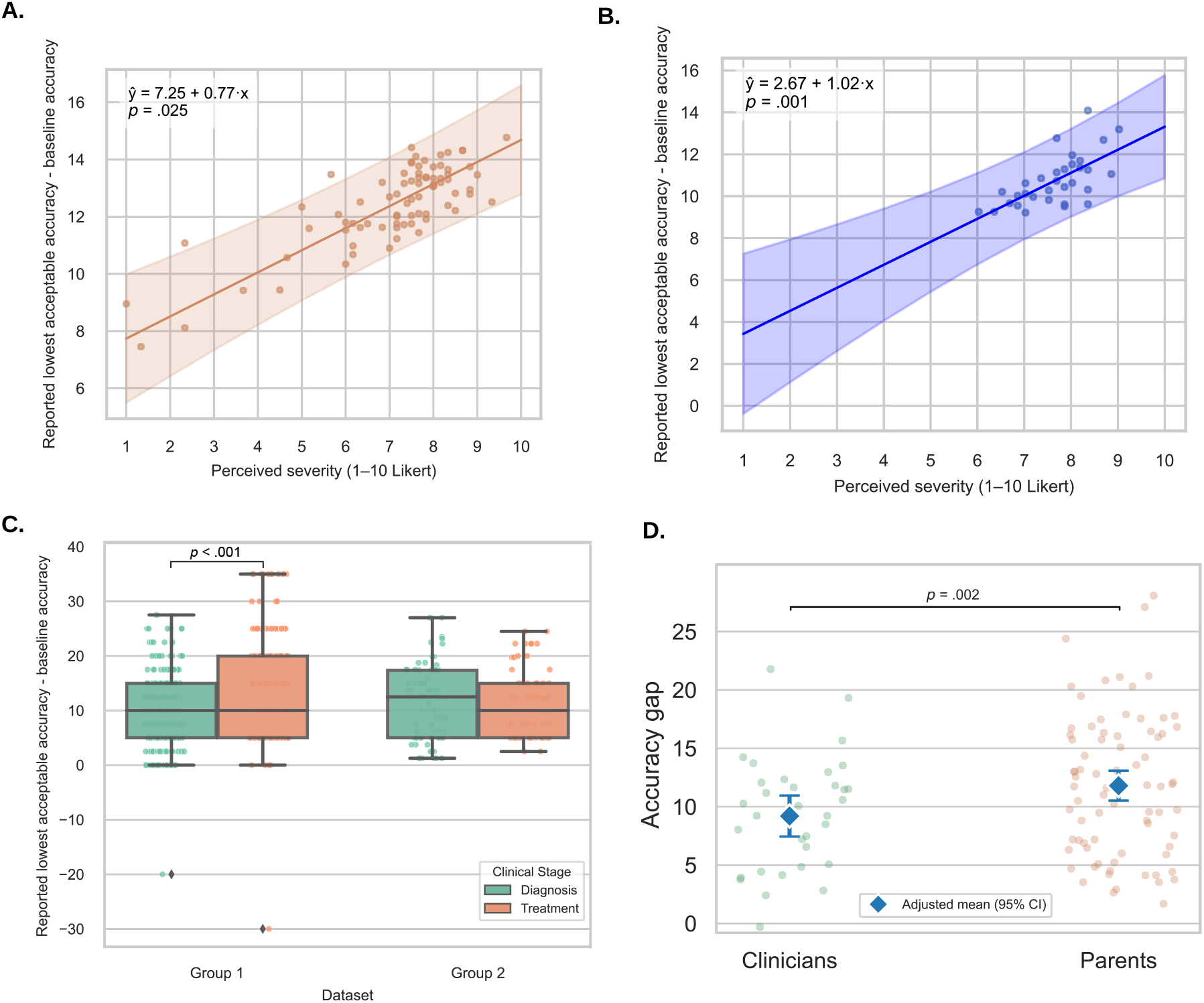
Effects on the lowest acceptable accuracy to use AI. (**A**) Difference between the lowest acceptable accuracy and baseline accuracy by application scenario (diagnosis vs. treatment) in the two parent groups. A significant difference was observed only in parent group 1. (**B**) Association between perceived disease severity and the lowest acceptable accuracy in parent group 2. Points represent participant-level responses adjusted for all other covariates included in the linear mixed-effect model. The solid line shows the model-predicted relationship, and the shaded area represents the 95% confidence intervals (CIs). (**C**) Association between perceived disease severity and the lowest acceptable accuracy in the clinician group. Points represent participant-level responses adjusted for all other covariates included in the linear mixed-effects model. The solid blue line shows the model-predicted relationship, and the shaded area represents the 95% CIs. (**D**) Difference in the accuracy gap between parent group 2 and clinicians. Jittered points represent participant-level responses adjusted for all other covariates included in the linear mixed-effects model. Blue diamonds indicate the estimated marginal group means, and error bars represent the corresponding 95% CIs.

### Maximal acceptable error rate to use AI

In the first parent group, higher digital usage was associated with greater tolerance of AI errors (Gamma GLM: *b* = 0.075, *p* = .037; Spearman’s *ρ* = .15, *p* = .029; Fig. 4A). This effect was not replicated in the second parent group. Instead, for the second parent group, higher levels of previous medical knowledge significantly predicted lower acceptable error rates (*β* = -.23, *p* = .030; see Fig. 4B). When utilizing the same minimal predictors as the first parent group, higher years of education in the second parent group predicted lower acceptable error rates (*β* = -.23, *p* = .025; see Fig. 4C). No significant effects of education years were found in the first parent group. No significant effects were observed in the clinical group, and acceptable error rates did not differ significantly between clinicians and the second parent group. Complete statistical parameters including t-statistics, degree of freedom (*df*), and 95% confidence intervals are listed in Supplementary Table S5.

**Fig. 4.**
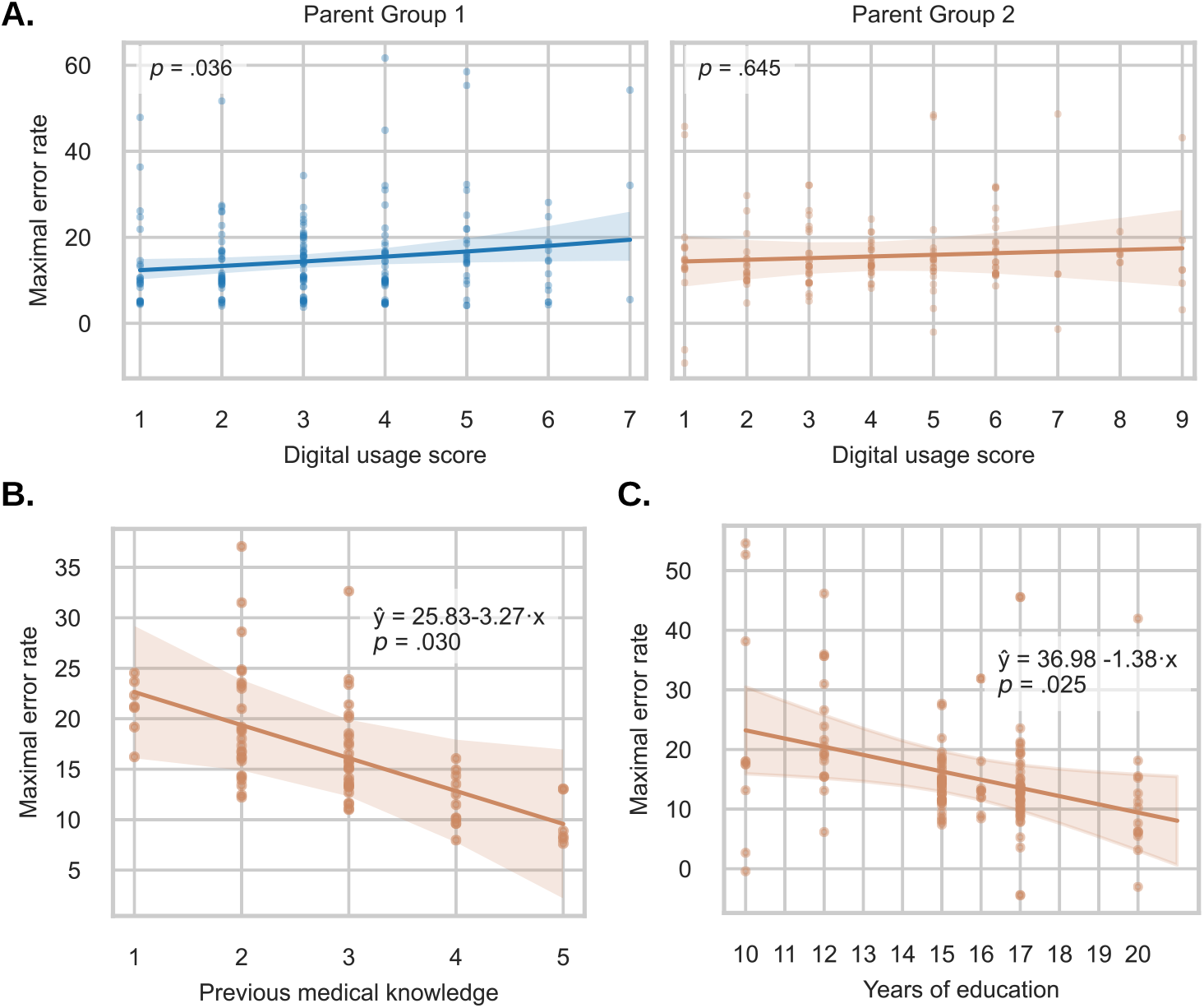
Effects on the maximal acceptable error rate. (**A**) Association between digital usage score and the maximal acceptable error rate in the two parent groups. A significant positive association was observed in parent group 1, whereas the effect was not replicated in parent group 2. Points represent participant-level responses adjusted for all other covariates included in the respective regression models. Solid lines indicate the model-predicted relationships, and shaded areas represent the 95% confidence intervals (CIs). The left panel shows predictions from the generalized linear model fitted to parent group 1, whereas the right panel shows predictions from the linear mixed-effects model fitted to parent group 2. (**B**) Association between previous medical knowledge and the maximal acceptable error rate in parent group 2. (**C**) Association between years of education and the maximal acceptable error rate in parent group 2 in the cross-sample analysis. Points represent participant-level responses adjusted for all other covariates included in the linear mixed-effects model. Solid lines indicate the model-predicted relationships, and shaded areas represent the 95% CIs.

### Willingness to use AI across privacy levels

Data privacy level robustly influenced willingness to use AI across all three groups (all *p* < .001; partial R^2^ = .215, .215, and .263 for parent group 1, parent group 2, and clinicians, respectively), with willingness declining markedly as data sharing became less restricted (see Fig. 5A). Furthermore, in the second parent group, perceived disease severity positively predicted willingness regardless of the privacy level (*p* < .001; see Fig. 5B). Post-hoc pairwise comparisons revealed critical privacy thresholds where acceptance dropped most significantly (Fig. 5A). For both parent groups, the largest declines occurred when data sharing moved from internal to external companies and when data left the EU borders (*p* ≤ .005). Clinicians showed more selective resistance, while they were tolerant of initial sharing expansions, they significantly rejected sharing with external organizations or international entities (*p* < .001).

Follow-up estimated marginal means comparisons indicated that males showed significantly higher willingness than females under higher data-sharing conditions (see Fig. 5C). Specifically, in the first parent group, males reported significantly higher willingness than females when data were shared with external companies within Germany (estimate = 1.32, SE = 0.52, *p* = .011, model-adjusted Hedges’s *g =* 0.61) and within the EU (estimate = 1.02, SE = 0.52, *p* = .050, model-adjusted Hedges’s *g =* 0.48). No significant gender differences were observed across any privacy level in the second parent group. In the clinician group, males also reported higher willingness than females when data were shared with external companies within the EU (estimate = 2.74, SE = 1.11, *p* = .019, model-adjusted Hedges’s *g =* 1.42). Detailed statistical results of LMM models and the interaction effects across privacy levels and groups are listed in Supplementary Table S6.

**Fig. 5.**
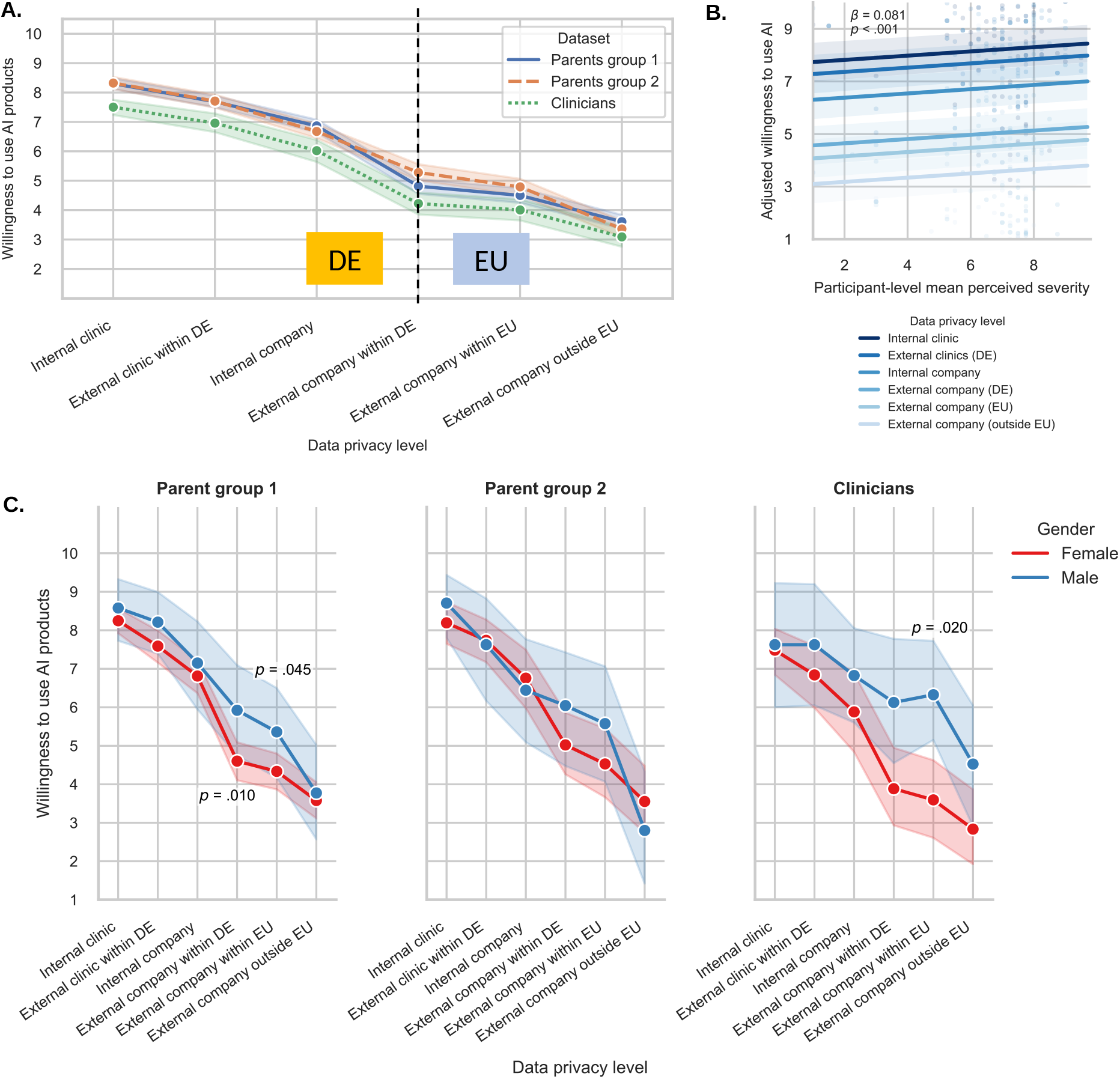
Effects on data privacy on willingness to use AI. (**A**) Willingness across privacy levels in the two parent groups and the clinician group. (**B**) Effect of perceived disease severity in the second parent group. Points represent participant-level adjusted willingness after controlling for covariates in the linear mixed-effects model. Colored lines represent the estimated marginal predictions for each privacy level, with shaded bands indicating 95% confidence intervals (CIs). (**C**) Willingness to use AI across the six data privacy levels by gender in the two parent groups and the clinician group. Lines represent group means, and the shaded areas indicate the 95% CIs. Significant gender differences based on estimated marginal means comparisons were observed at data-sharing levels involving external companies within Germany and the European Union in parent group 1, and at the European Union level in the clinician group.

### Decisions during AI-Clinician inconsistency

When faced with a conflict between AI and clinician recommendations, participants across all groups were least likely to choose AI over a clinician (see Fig. 6A). Instead, the most frequent choice was to consider both options but ultimately choose the clinician’s recommendation. However, participants were consistently more likely to trust the AI’s recommendation in treatment scenarios than in diagnostic ones across the first parent group (odds ratios, *OR* = 2.35), the second parent group (*OR* = 4.17) and clinicians (*OR* = 7.78; see Fig. 6B).

In the parent groups, higher digital usage was associated with increased odds of trusting AI (*OR* = 4.04) or considering it as a viable alternative to clinicians (*OR* = 1.77; see Fig. 6C). For the second parent group, higher levels of previous AI (*OR* = 1.58) and medical knowledge (*OR* = 1.48) further increased the likelihood of choosing AI. Conversely, higher family income (*OR* = 0.68) and more years of education (*OR* = 0.82) were associated with a preference for clinician-led decisions (see Fig. 6D).

Among clinicians, trust in AI also increased with digital usage (*OR* = 1.66), while perceived disease severity was associated with increased odds of considering both options but ultimately following their own judgment (*OR* = 1.43; see Fig. 6E). These patterns remained robust when modeling parents and clinicians together (*OR* = 1.27 for digital usage), with no significant differences found between the two groups. Detailed statistical parameters including OR and 95% credible intervals (Crl) are listed in Supplementary Table S7.

**Fig. 6.**
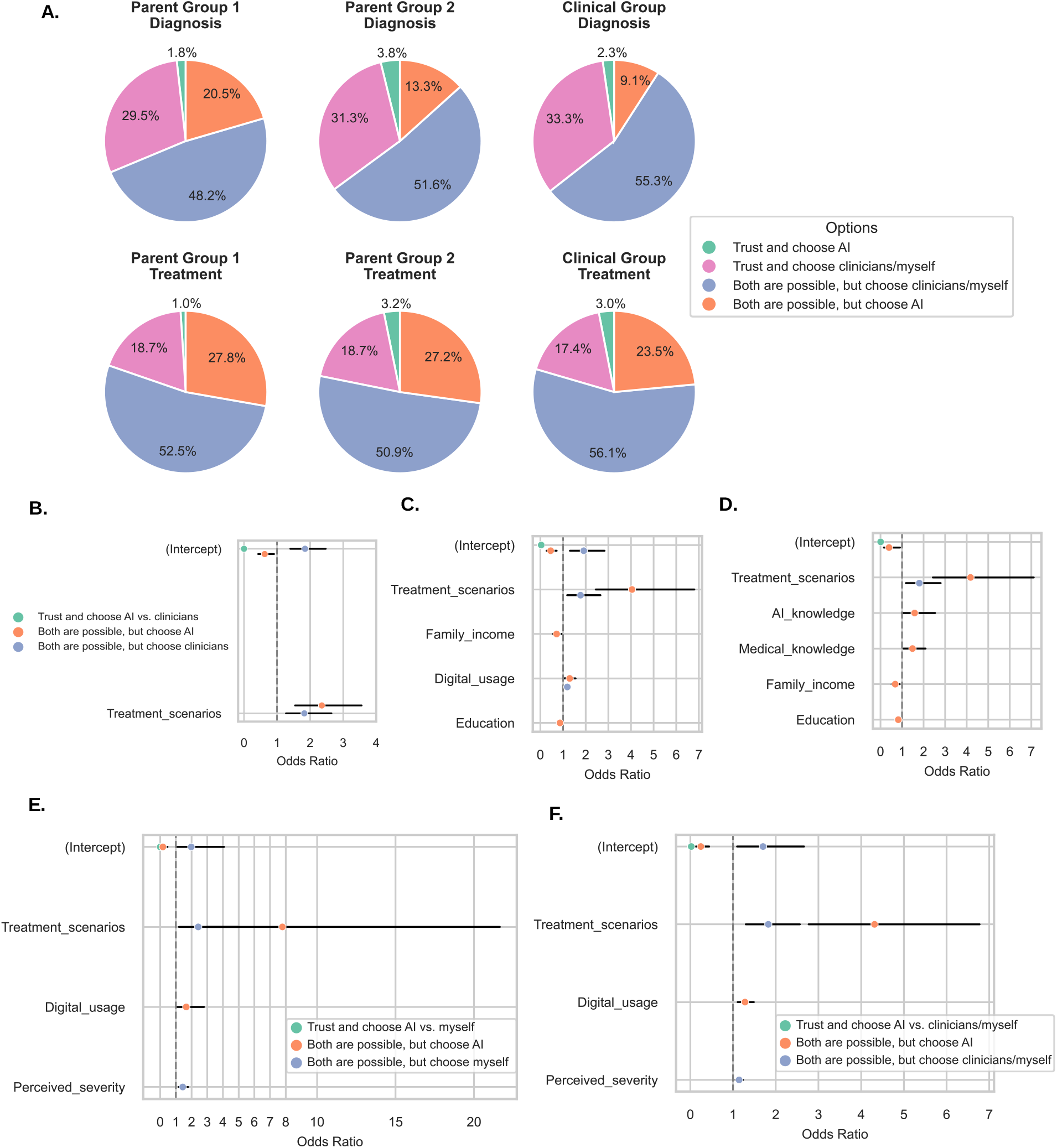
Decisions during AI-clinician inconsistency. (**A**) Distribution of decision categories by application scenario (diagnosis vs. treatment) and participant group. (**B-F**) Odds ratios (ORs) from Bayesian multilevel categorical models, with 95% credible intervals (CrIs) shown as black horizontal error bars. ORs greater than 1 indicate increased probability of choosing the corresponding response category relative to the reference category (“trust and choose clinicians/myself”), whereas ORs less than 1 indicate decreased probability. (B) Parent group 1. (**C**) Parent group 2 (replication analysis). (**D**) Parent group 2 with the full predictor set. (**E**) Clinician group. (**F**) Combined analysis of parent group 2 and the clinician group.

## Discussion

Overall, our findings show that acceptance of AI in pediatric healthcare is shaped primarily by clinical context, data privacy, and the role of human judgment. Replicated and new findings indicate that perceived disease severity, digital experience, and previous knowledge further influence acceptance towards AI applications, while differences between parents and clinicians suggest that AI implementation and communication should be tailored to stakeholder needs.

### Replicated and convergent findings across groups

Across parents and clinicians, willingness to use AI in pediatric healthcare depended strongly on the clinical context. The acceptance of AI applications was consistently highest for severe conditions such as cancer and lowest for developmental-language applications. This pattern suggests that perceived clinical benefit may promote AI acceptance in high-stakes contexts, while applications involving developmental assessment and sensitive pediatric data may evoke greater caution among parents. These findings highlight a context-dependent acceptance of AI in pediatric settings.

A second convergent finding was the consistent decline in willingness as the scope of data sharing expanded. Willingness to use AI decreased as data sharing moved from internal clinical use to external organizations, with particularly large declines when children’s data were shared with companies or outside the EU. This pattern was evident in parents and clinicians and is consistent with ethical concerns about non-maleficence, data control, and the protection of children’s information (23–25). At the same time, higher perceived disease severity consistently increased parental willingness across privacy levels, suggesting that perceived clinical benefit may partly offset concerns about data sharing. Parents may accept broader data use when they believe it could substantially improve care, but this does not remove the need for transparent consent procedures, clear data governance, and ongoing control over how data are used (26).

Participant responses to AI–clinician disagreements converged across groups: participants preferred clinician-led decision-making and least trusted AI when it contradicted professional judgment. However, AI recommendations were more acceptable in treatment than in diagnostic scenarios. This caution stems from the critical need for diagnostic accuracy, where missed or incorrect diagnoses carry severe downstream consequences (e.g. ineffective interventions). Nevertheless, AI provides distinct value as a diagnostic support tool in complex pediatric cases with unexplained symptoms, notably by generating hypotheses for rare conditions and evaluating differential diagnoses. Positioning AI as a complementary tool that augments rather than replaces clinical judgment could address parental concerns and increase parental acceptance while maintaining necessary human oversight (27–29).

### Non-replicated and new findings in the second parent group

The preference for treatment-related AI applications observed in the first parent group was not replicated in the second parent group. The first questionnaire linked diagnosis and treatment to different diseases (e.g., diagnosis AI for pneumonia, treatment AI for cancer), whereas the second questionnaire assessed all diseases in both application scenarios. The original effects of application scenarios may therefore have been partly confounded by disease type. The second cohort instead showed that perceived disease severity was positively related to willingness and accuracy requirements. Parents were more willing to use AI for their children in severe conditions, such as cancer, while requiring a larger accuracy advantage over conventional care.

The overall gender difference observed in the first parent group was not replicated in the second parent group, suggesting that lower willingness among women may not represent a general pattern. Instead, gender differences appeared more selectively in relation to data privacy. In the first parent group, women were less willing than men when their children’s data were shared with external companies within Germany or the EU. However, these contrasts should be interpreted cautiously given their cohort-specific nature and the gender imbalance in both parent samples.

The second parent group provided additional evidence that family and knowledge-related factors shape AI acceptance. Parents with older children reported greater willingness, possibly because accumulated experience with healthcare decisions supports more confident evaluation of new tools

(30). Greater perceived disease severity predicted willingness to use AI, whereas previous medical knowledge and years of education were associated with greater tolerance of AI errors. These results suggest that AI acceptance depends not only on perceived clinical need, but also on the ability to interpret clinical uncertainty and potential trade-offs. Communication should therefore address both the expected clinical benefit of AI and its performance limitations in a clear and context-specific manner.

### Clinician-specific findings

Among clinicians, digital engagement was particularly relevant. Clinicians who used more digital products in daily life were more willing to adopt AI, and their willingness increased more strongly with digital usage compared to parents. This is consistent with evidence that technology exposure and AI training facilitate professional adoption (31,32). Targeted training should therefore combine practical experience with information about AI limitations, appropriate use, and clinical accountability rather than focusing only on technical functionality.

Clinicians also appeared more tolerant of imperfect AI performance than parents. They required a smaller improvement over baseline accuracy to accept using AI and were more willing to choose AI in treatment scenarios. Familiarity with diagnostic uncertainty and the fallibility of conventional care may help clinicians interpret imperfect performance more realistically (33–35). However, as disease severity increased, clinicians became more likely to rely on their own judgment rather than AI. This may reflect heightened concerns about responsibility and liability in high-risk cases, highlighting that professional adoption does not imply willingness to surrender clinical authority (35–40).

### Comparison between parents and clinicians

Direct comparisons revealed both shared concerns and context-specific differences between parents and clinicians. Willingness declined similarly across groups as data sharing expanded, indicating that privacy is a shared concern across parents and clinicians in AI usage. Both groups also preferred human oversight when AI and clinicians disagreed. However, group differences in willingness were disease-specific rather than systematic. Specifically, clinicians reported lowered willingness for asthma and cancer whereas two parent groups were comparable across diseases. Parents may evaluate AI primarily in terms of potential consequences for their child, whereas clinicians may draw more on professional knowledge and experience with uncertainty, competing risks, and imperfect clinical alternatives.

These differences have practical implications for communication. Parents may benefit from clear explanations of why a given accuracy level is clinically meaningful, what happens when AI fails, and who remains responsible for the final decision. Clinicians may benefit more from workflow-specific evidence, hands-on training, and explicit guidance on human oversight towards AI. Therefore, communication strategies should be tailored to stakeholder roles rather than assuming that a single approach will build trust across groups.

### Ethical and regulatory implications

To sum up, the current results support three priorities for pediatric AI: proportional accuracy requirements, strong privacy governance, and preserved human oversight. The EU AI Act requires high-risk systems to meet appropriate standards of accuracy, robustness, transparency, data governance, and human oversight (12). These requirements are particularly important for AI applications involving the developmental data of children and adolescents, given the relatively low levels of acceptance observed in our results and the sensitive nature of such data. AI providers should disclose known limitations and potential failure modes, clearly specify how and where data are stored, processed, and shared, and provide users with informed consent and control. Trust is more likely to develop when safeguards are concrete, transparent, and understandable rather than presented as general assurances.

### Limitations

#### Questionnaire design and study methodology

The study measured self-reported willingness using hypothesized clinical scenarios. Acceptance may change when families and clinicians encounter AI in real clinical settings due to intention-behavior gaps (41,42). Additionally, the two questionnaires developed for two parent groups were not fully identical, particularly in how diseases were assigned to diagnosis and treatment, limiting direct replication of some effects. Finally, the structured questionnaire could not capture the detailed reasoning behind individual choices. Longitudinal and qualitative studies (e.g., semi-structured interview) should examine whether these initial acceptances translate into sustained use and how responsibility, privacy, and AI performance are negotiated in practice.

#### Sample characteristics and generalizability

Compared to the first parent group, the sample sizes of the second parent group and clinician group were relatively small, reducing statistical power for sub-group and cross-sample analyses. Women were overrepresented across all three groups, which may have limited the possibility of detecting gender effects. Moreover, some clinicians were also parents, which may have influenced their responses despite instructions to answer from a professional perspective. Future research should account for the parental status of clinicians as a covariate to prevent potential bias brought from the dual identity. Furthermore, children and adolescents themselves were not included as participants in the current study. Although parents and healthcare professionals represent key stakeholders in pediatric healthcare and often play important roles in healthcare decision-making and the implementation of AI, pediatric healthcare ultimately concerns children and adolescents. Their perspectives may differ from those of adults, particularly among adolescents, whose autonomy and involvement in healthcare decisions increase with age (43,44). Future research should evaluate their specific acceptability, expectations, and concerns regarding AI.

## Conclusions

Our study demonstrates that, given the explicit choice, both parents and clinicians prefer to trust the expertise of clinicians over AI, although they consistently rate their willingness to use AI at a relatively high level. However, the acceptance is context-dependent, with both parents and clinicians showing a significant preference for AI in treatment over diagnosis scenarios. Individuals expect AI to offer an accuracy boost, aligning with the ethical principle of beneficence. Notably, perceived disease severity plays a positive role in driving parents’ willingness to use AI and their acceptable accuracy threshold. Higher digital usage and prior medical knowledge smooth the path toward AI acceptance by increasing error tolerance. As data sharing levels increase, especially when extended to the national and EU level, willingness to use AI largely decreases. Interestingly, as perceived disease severity increases, clinicians are more likely to prioritize their own judgment over AI during inconsistent conclusions. To sum up, implementing digital training, bridging knowledge gaps in AI and the medical field, building ethical frameworks that ensure data privacy, and establishing distributed responsibility could facilitate the acceptability of AI in clinical care among parents and clinical staff.

## Data Availability

All data produced in the present study are available upon reasonable request to the authors.

## Notes

### Competing Interest Statement

The authors have declared no competing interest.

### Author Declarations

Ethics committee of the Medical Faculty of Heinrich-Heine-University Duesseldorf gave ethical approval for this work.

### Summary of Updates

An author's name was mis-typed; two authors' ORCID were added.

### Funding

- German Federal Ministry of Education and Research (BMBF): 01GP2203A

